# Global pattern of COVID-19 research

**DOI:** 10.1101/2020.07.04.20146530

**Authors:** Xunzhi Zhu, Qi Jin, Xiyi Jiang, Yuanyuan Dan, Aimin Zhang, Guangming Qiu, Jianlin Lou, Hualong Yu

## Abstract

Since the COVID-19 outbreak began, a large number of studies have been conducted in a short period. However, it is unclear whether countries involved in this crisis have made adequate efforts and allocated resources to cutting-edge SARS-CoV-2 research. We analyzed the dynamics of and professional fields represented by papers about this novel coronavirus published before June 15, 2020. High-infection countries produced more scientific output than low-infection countries, and high-income and upper-middle-income countries were the main contributors. However, the research areas overlapped substantially, indicating a waste of resources. Our findings also suggest that international cooperation among countries is still relatively lacking, and all countries should make better use of their strengths to face the epidemic jointly.

## Main Text

The human being is no stranger to the threat of infectious diseases. Over the human history of battling diseases, scientific researchers have played a key role in disease prevention and control. For instance, smallpox was eliminated successfully with the cowpox vaccine, developed by Edward Jenner, and cholera would have proliferated if John Snow had not discovered, through his remarkable epidemiological research, that the pathogen spread by contaminated water (*1, 2*). Knowledge about the characteristics of Coronavirus Disease 2019 (COVID-19) is accumulating with the rapid response of the scientific community, and considerable progress has been made. The genome sequence of the virus was first identified on January 10, 2020 by a Chinese research team shortly after a cluster of unrecognized pneumonia was first reported in China (*3, 4*). The identification of the genome sequence enabled different laboratories to design diagnostic tests to precisely detect the viral RNA, and it facilitated the development of antiviral drugs and vaccines (*5, 6*). Various diagnostic methods have been applied, which has accelerated the detection of infected people (*7*). Importantly, early characterization of virus transmission dynamics and patients’ clinical characteristics typically provide a solid basis for governments’ deployment of prompt public health measures, such as intensive surveillance and social distancing, and mobilization of health resources (*8, 9*). However, while various scientific efforts are warranted, many urgent scientific questions remain unanswered. For example, although bats and pangolins were suspected animal hosts of SARS-CoV-2, no consensus on the origin of the disease has been reached for the implementation of defensive barriers against reemergence (*10*). Multiple scientific efforts are underway to devise new vaccines with ongoing clinical trials, but no specific therapeutic agents or preventive vaccines for the COVID-19 are currently approved, and it is anticipated that a vaccine will not be available before 2021 (*11, 12*).

Since a cluster of cases was first reported in December 2019, the disease has spread across borders at unexpected speeds and was declared a pandemic in March 2020, owing to its intercontinental transmission (*4*). As of June 15, 2020, the disease had spread to 200 countries with over 7,823,289 confirmed cases, including 431,541 deaths (*13*). In addition to the widespread public health threat, the disruption experienced in economics, trade, education, and other social domains has been devastating and may induce long-term consequences (*14, 15*). Combatting COVID-19 requires close collaboration between countries, especially among their scientific communities. Countries vary in their overall scientific research strengths, but each may offer advantages in a specific research field that can complement their counterparts. Thus, collaborative work is assumed to be more efficient than isolated work (*16*). When facing this urgent global crisis, it is essential to mobilize and optimize available scientific and social support to maximize research efficiency and avoid redundancy. Therefore, the aim of this study was to assess the global pattern of COVID-19 research and provide a reference for optimizing scientific resources. To that end, we conducted a bibliometric analysis of COVID-19-related researches, reviewed scientific contributions of several representative countries, and evaluated the extent of international cooperation.

We collected all COVID-19-related research published before June 15, 2020 from PubMed. First, we explored the global temporal dynamics of published research. Using a log-linear model, we tested the relationship between research strength and severity of the epidemic in countries all over the world. Research strength was determined by number, types, and distribution of publications, the combination of which was assumed to be a reflection of a country’s research efforts and prowess. Second, we divided the published papers into the following ten categories: A. Origin and gene sequencing; B. Epidemiological characteristics; C. Clinical characteristics; D. Pathology and molecular biology; E. Diagnostic methods; F. Treatment; G. Vaccine development; H. Prevention and control strategies; I. Studies targeting a specific population; and J. Reviews. These categories represent the main fields involved in novel coronavirus research. Lastly, to evaluate the global collaborative situation, we analyzed the joint publications of authors from multiple countries; we believe that international cooperation is essential for fighting this new pandemic.

Between January 21, 2020, when the first research article on the novel coronavirus was published, and June 15, 2020, 11,859 original papers from 157 countries were posted (papers with no country or area information were excluded). Overall, the number of global publications increased over time. The growth curve comprised three parts: a 2-month lag phase, a 2-month exponential growth phase, and a period of constant rise (**Fig. 1a**). The 2-month exponential growth phase suggests that as the pandemic spread, accumulating input resources led to increased research productivity. Twenty countries contributed the most of publications (**Fig. 1c**). The US, China, Italy, and the UK contributed the most papers (**Fig. 1d**). Of these, the US produced the most studies with 3,077 publications (26.9%), followed by China with 1,566 releases (16.9%) (**Fig. 1d**). It is not surprising that the majority of papers were attributed to US institutions because the US has the most abundant scientific resources. Furthermore, it currently reports the largest number of confirmed COVID-19 cases and related deaths, which may help explain why so many of those resources have been devoted to preventing further spread of SARS-CoV-2 (*17*). Indeed, this pattern persisted for most of 20 specified countries, although the time began to increase, and the total number of publications varied (**Fig. S1**).

**Fig. 1.**
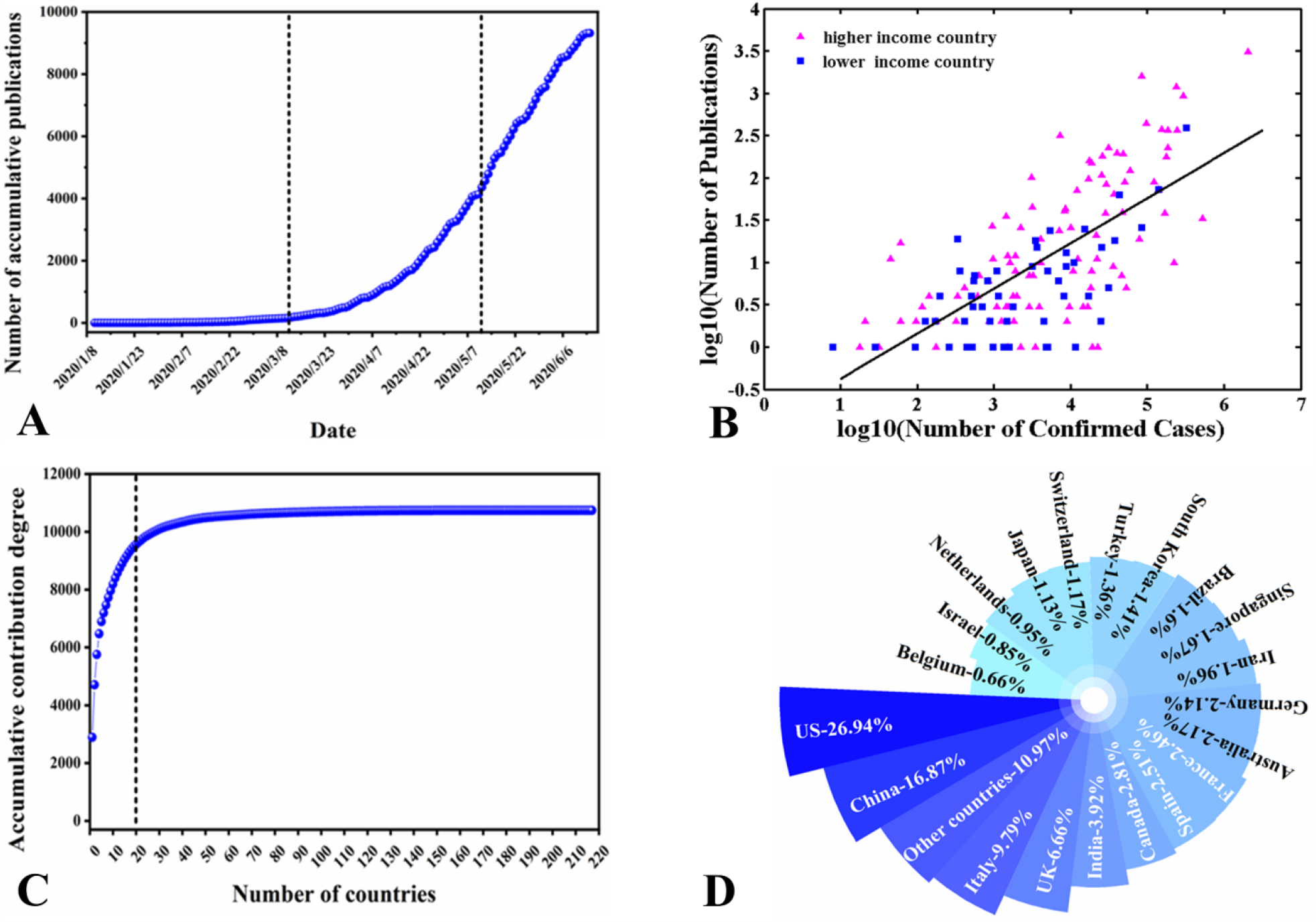
Relationships between publication count and (A) time, (B) reported cases, and (C and D) nations. (A) Publications about COVID-19 increased in three stages: a lag phase, an exponential growth phase, and a period of constant rise. (B) Violet dots represent higher-income countries, and blue dots represent lower-income countries. The line indicates a significant fitted regression with *P*<0.001. (C, D) These 20 countries are the main contributors to scientific publications.

The relationship between the number of confirmed cases and the publication count in each country was also described (**Fig. 1b**). Figure 1b shows an overall upward linear pattern (R^2^=0.48, *P*<0.001), indicating that the number of publications increased with the number of cases. An increase in confirmed cases represents the spread of infection; this could be a driving force behind research initiation even as it provides sources (e.g., participants) for research. The pattern persisted for both higher-income groups (high- and upper-middle-income countries) and lower-income groups (low- and middle-income countries). Compared with higher-income groups, lower-income groups had fewer publications and COVID-19 cases. This is reasonable because high-income countries generally exhibit superior research strength and can more easily get support from their human resources, which contributes to more research activity. Interestingly, several countries reported case numbers that varied from single digits to more than 100,000 yet evidenced no publications.

In terms of research focus, the predominant publication categories were: diagnostic methods (36.4%), followed by studies targeting a specific population (18.7%), and prevention and control strategies (10.3%) (**Fig. 2c**). Publications about vaccine development tended to be in the minority (0.96%) (**Fig. 2c**), which may be mainly attributable to the lengthy vaccine-development process using randomized control trials (RCTs) (*18*). Publications from the US, China, Italy, and the UK were content-rich, with distribution across ten research domains, suggesting that more resources had been devoted to scientific research in these countries in the previous few months (**Fig. 2a**). Generally, in all observed nations, research on diagnostic methods received the most attention, which was consistent with the research outputs, especially for the US (n=1,621), followed by China (n=1,293). This indicates the difficulty and importance of accurately diagnosing COVID-19 and a low research threshold in diagnostic methods. Notably, publications from China also focused on epidemiological characteristics (n=318) and clinical characteristics (n= 357), which play essential roles in pandemic/epidemic prevention and control. The US produced more publications about vaccine development compared with other countries, although the number was still small (n= 66). Remarkably, the research hotspots have remained stable in each of the ten categories (**Fig. S2)**. To date, no specific antiviral treatment has been confirmed to be effective against COVID-19, which emphasizes the importance of developing a targeted vaccine for the general population (*19*). Therefore, increased focus on vaccine development is urgently needed at this stage.

**Fig. 2.**
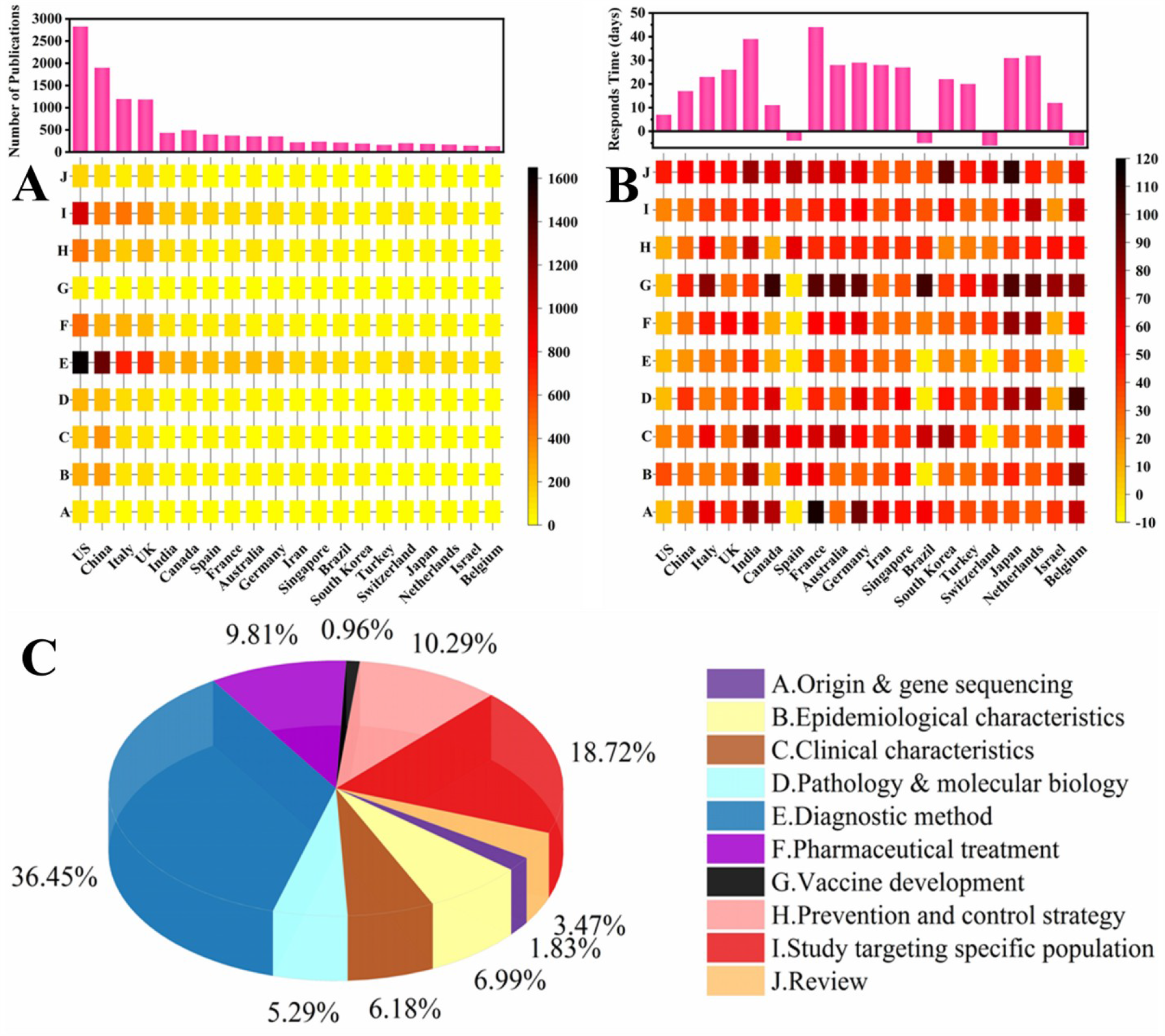
Publication patterns of COVID-19 research by (A) quantity and (B) response time in ten research fields. (C) Developmental imbalances exist in the ten reviewed fields.

In terms of response time, Belgium and Spain were the earliest, and their first papers were published before the first case in their country was confirmed (**Fig. 2b**). Switzerland and Brazil also had publications out before the first case was reported in their respective countries, but the absolute time was longer. Further stratification by research focus demonstrated that diagnostic methods received the earliest (4–6 days ahead COVID-19 outbreak in their country) attention across all nations, especially in Belgium, Brazil, Spain, and Switzerland, which was also reflected in their research outputs. However, most countries focused on vaccine development later, which coincided with the focus on diagnosis and treatment during the initial outbreak. Notably, the UK published the first paper based on vaccine development. To summarize, all the 20 countries responded quickly to the COVID-19 pandemic.

More than two-thirds of the reviewed studies were conducted independently, and among collaborative studies, the majority was authored by scientists from two or three countries (**Fig. 3b**). The most frequent cooperation was between the US and China, the two most significant contributors to COVID-19-related research (**Fig. 3a**). Strong scientific collaborations were also apparent among the US, China, the UK, and Italy (**Fig. 3a**). These four countries also lead in COVID-19 research. Of the ten research focus categories, studies targeting a specific population showed the most extensive international collaboration; all 20 countries produced joint publications (**Fig. S3**). Less comprehensive, but reliable enough, cooperation was detected in the category of treatment; most countries conducted collaborative work in this area (**Fig. S3**). Diagnostic methods, the largest publication category, also witnessed frequent and broad cross-border collaboration (**Fig. S3**). In contrast, the categories origin and gene sequencing, and vaccine development were more likely to be authored individual contributors. This difference may be attributable to the particular challenges and commercial hurdles faced in vaccine development; most countries collaborating in vaccine development are recognized as developed countries with high research capability. Generally, vaccine development requires other studies as a basis. Currently, there is relatively little published vaccine research, and therefore it is possible that there is a time lag in this field. Furthermore, western developed countries conducted more collaborative work than eastern countries, and the pattern is more apparent when presented on a relative scale (**Fig. 3c**). The most substantial proportion of collaborative work was seen in Switzerland, the Netherlands, Israel, and Belgium, accounting for above 75% of national publications. Western countries are generally more active in scientific collaboration. As to the collaboration pattern of COVID-19 research, most research is purely domestic in eastern countries,, and international collaborative work contributes to less than 40% of all publications in China, India, Iran, and Korea.

**Fig. 3.**
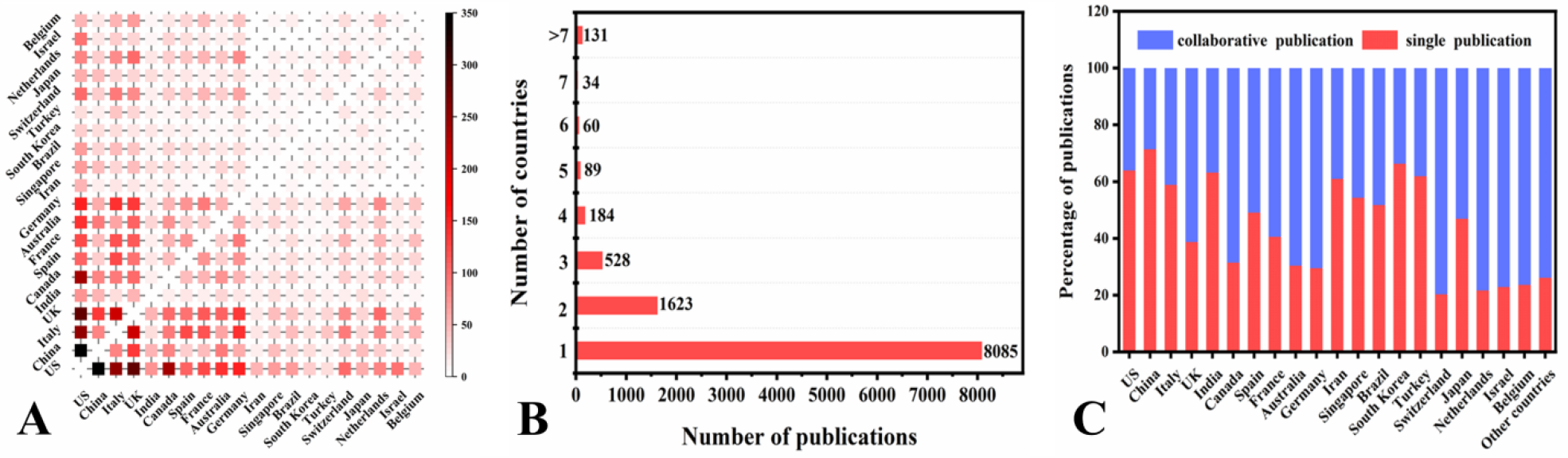
Collaboration status of the main contributing countries in COVID-19 research (A to C). (A) More collaborations are represented by darker colors in the heatmap. (B) Most of the research was produced by one country. (C) Red bars represent the proportion of solo-authored papers; blue bars represent the proportion of collaborated papers.

International collaboration promotes the sharing of ideas, resources, and outcomes among research teams, and it is likely to yield high-quality output (*19, 20*). The cost of international collaboration has decreased largely because of globalization and the development of information technology. Many countries have established specialized centers to facilitate high-quality science and technological innovation through collaboration (*21*). However, in this analysis, we found inequities in cooperation between western and eastern countries. This finding is consistent with a previous study, which reported that in established economies such as the US and Europe, international collaboration accounted for a growing percentage of annual research output over decades, while in emerging economies like China, India, and Brazil, domestic research remains the predominant research mode (*16*). Such differences may be attributable to variations in geographical position, cultural background, and political system.

Since its discovery, COVID-19 spread quickly and has caused enormous economic loss and human casualties in a short time. With rapid developments in science and technology, personal preparedness for emerging infectious diseases has improved substantially compared with past centuries. Our study suggests that, while response speeds varied across countries, the world scientific community responded quickly to the new disease, which has led to significant achievements in diverse research fields. However, more collaboration among nations is needed. Similar research patterns in different countries suggest homogeneous competition occurred, which was, arguably, a waste of public resources. Countries should draw on one another’s strengths, try to reduce research duplication, and strengthen international divisions of labor and cooperation while giving full play to their respective strengths. Higher-income countries, whose economic advantages offer technological advantages, could help lower-income countries. The present tragic and unprecedented pandemic has involved almost the entire human race, yet it also presents an excellent opportunity for the world to unite.

## Data Availability

All data are available in the main text or the supplementary materials.

## Acknowledgments

We thank Anita Harman, PhD, from Liwen Bianji, Edanz Editing China (www.liwenbianji.cn/ac), for editing the English text of a draft of this manuscript.

## Author contributions

X.Z., H.Y., and J.L. designed the study. H.Y., A.Z., and G.Q. collected the data. X.Z., H.Y., A.Z., and G.Q. performed statistical analysis. X.Z., Q.J., X.J., H.Y., J.L., and Y.D. wrote the draft of the manuscript. All authors contributed to revisions and gave final approval for publication;

## Data and materials availability

All data are available in the main text or the supplementary materials.

## Supplementary Materials

Methods

Figures S1-S3

Table S1

## Supplementary Materials

### Methods

#### Data collection

We collected the data from the PubMed database (https://pubmed.ncbi.nlm.nih.gov/), which contains the most comprehensive list of scientific publications in medicine and life sciences. The data comprise the following publication information: *Title, Author Affiliations, Journal Name, Publication Date* and *Keywords*. The search terms were: “novel coronavirus 2019,” “coronavirus 2019,” “COVID 2019,” and “COVID-19.” The publication language was English. The publication date range was restricted to 1st December, 2019 to 15th June, 2020. Additionally, we filtered publication type, thereby excluding *News, Comments, Erratums, Letters*, and *Editorials*.

We retrieved 11,859 records that satisfied all search conditions listed above. Subsequently, duplicate records were removed from the results by simultaneously comparing the *Title* and *Journal Name* terms. Thus, when two or more records had exactly the same results for these two search terms, only one record was retained. After removing duplicate records, 11,721 records remained and were used for analysis.

### Publication information statistics and association with confirmed cases

First, we collected the standard name of each country or region in the world from the World Bank website (https://databank.worldbank.org/), which also puts all countries and regions into four non-overlapping groups, namely *high income, upper-middle income, lower-middle income*, and *low income*. By matching this information with the *Affiliation* search term results in our publication list, we extracted publication country information for each publication record. Some publication records were removed from our list at this point if their *Affiliation* information was missing; this left 10,738 records for further analysis.

Given that a research paper can be collaboratively finished by institutions in multiple countries, an indicator called *contribution degree* was defined. Suppose a paper is written by authors from *n* countries, then each country acquires a contribution degree of 1/*n*. After ranking the countries according to their contribution degrees in descending sort, a saturation curve was generated by associating the countries and their number of publications (see Fig. 1c). From the saturation curve, we extracted the top 20 contributing countries; in descending order, they are: *US, China, Italy, UK, Canada, India, France, Germany, Spain, Australia, Switzerland, Iran, Singapore, Netherlands, Turkey, Brazil, Japan, South Korea, Israel*, and *Belgium*. Fig. 1d shows the percentage of contribution degree for each one of these countries, as well as the combined contribution of the remaining countries. As the figure shows, the sum of contribution degrees belonging to the top 20 countries is 89.03%, which greatly exceeds that of the remaining 197 countries or regions.

Next, we investigated whether the association between the number of confirmed cases and the number of publications for the major countries. From the World Health Organization (WHO) website (https://covid19.who.int/), we determined the number of accumulated confirmed cases in each country or region up to 15th June, 2020. We were also interested in whether the high- and upper-middle income countries had contributed more to research on the novel coronavirus. Therefore, we divided all countries and regions into two new groups—higher income group (high- and upper-middle income countries) and lower income group (lower-middle and low-income countries). Fig. 1b shows the mapping relationship between the number of confirmed novel coronavirus cases and the number of publications in each country or region in a log scale. The acquired correlation coefficients *R*^2^ and the corresponding significance level *P* were used to observe the association strength between the number of confirmed cases and the number of publications.

Another basic statistic we measured was the association between the number of publications and the publication date, which can indicate the trend of research strength. This was achieved by checking the *Publication Date* search term results in our publication list; the research strength curve over time is presented in Fig. 1a. Publication records (20.46%) that lacked exact publication date information were excluded from analysis, and the remaining records were used to generate the research strength curve over time. Figure S1 presents the association between the number of publications and the publication date for each one in the top 20 countries to observe whether the different countries have the different responds for the novel coronavirus research.

#### Topic division, research strength and response time analysis

The existing publications related to the novel coronavirus cover many different topics. In this study, we focused on the following ten topics: A. *Origin and gene sequencing*, B. *Epidemiological characteristics*, C. *Clinical characteristics*, D. *Pathology and molecular biology*, E. *Diagnostic methods*, F. *Pharmaceutical treatments*, G. *Vaccine development*, H. *Prevention and control strategies*, I. *Studies targeting a specific population*, and J. *Review*. For each topic, some matching words, which were used to match the publication with a topic, had been collected in accordance with the advice of two medical experts (see Table S1). Matching words were used to match both *Title* and *Keywords* term results to indicate which topic or topics a publication belongs to.

After comparing the matching words with both *Journal Title* and *Keywords* search term results in our publication record list, 9,103 publication records were extracted. Then, the proportion of each topic was calculated (see Fig. 2c) to determine its current research strength or deficiency. Furthermore, topic information was associated with the *Affiliation* search term results to help assess the research strength of each top 20 (i.e., most publications) country for each topic and for all ten topics (see Fig. 2a). Additionally, the date of the first confirmed COVID-19 case in each of the top 20 countries was acquired from the WHO website (https://covid19.who.int/). By comparing the date of the first publication in each topic area and all topics in each country with the date of its first confirmed case, we could observe the response speed of each country and its research strengths and shortcomings (see Fig. 2b).

After checking the *Publication Date* search term results in our publication record list, we produced a research strength curve for each topic across time; this is presented in Fig. S2. This figure illustrates both response time and research strength for each topic, information that can further guide the scientists in making informed decisions about difficult topics.

#### Cooperation strength analysis

In this study, we also evaluated the strength of cooperation in novel coronavirus research among different countries. On the basis of the publication country information of each publication, we present the statistical results for the cooperation strength between any two top 20 countries (see Fig. 3a), the number of publications written by different groups of collaborative countries or regions (see Fig. 3b), and the proportion of publishing collaboration for each top 20 country (see Fig. 3c).

Finally, we also associated topic information with publication country information to observe whether some top 20 countries prefer to cooperate with each other on some topics but not on others (see Fig. S3). This helped us find the current collaborative shortcomings among different countries, which can further guide scientists and governments of different countries and regions to make better decisions about collaboration in future research on the novel coronavirus.

**Fig. S1.**
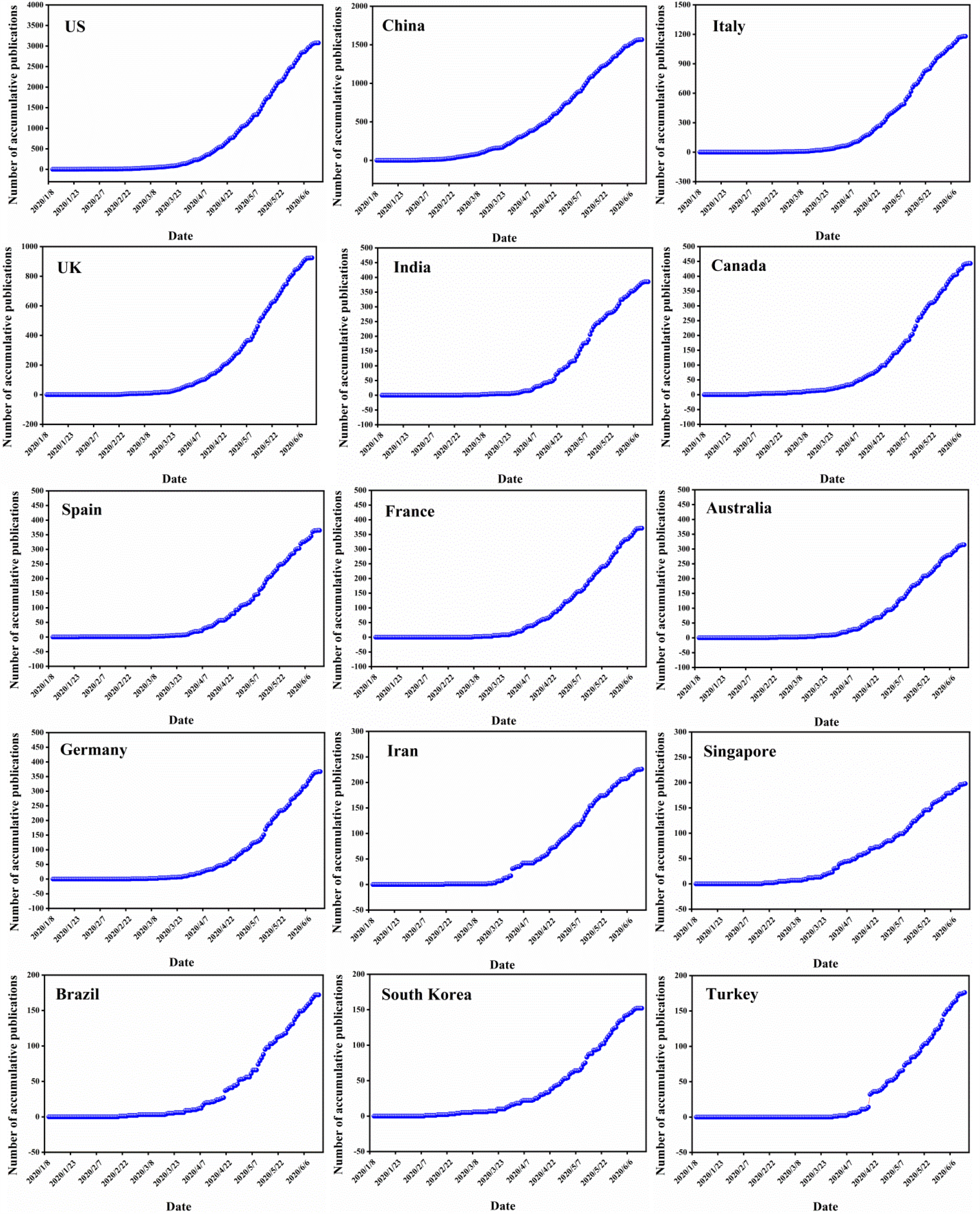
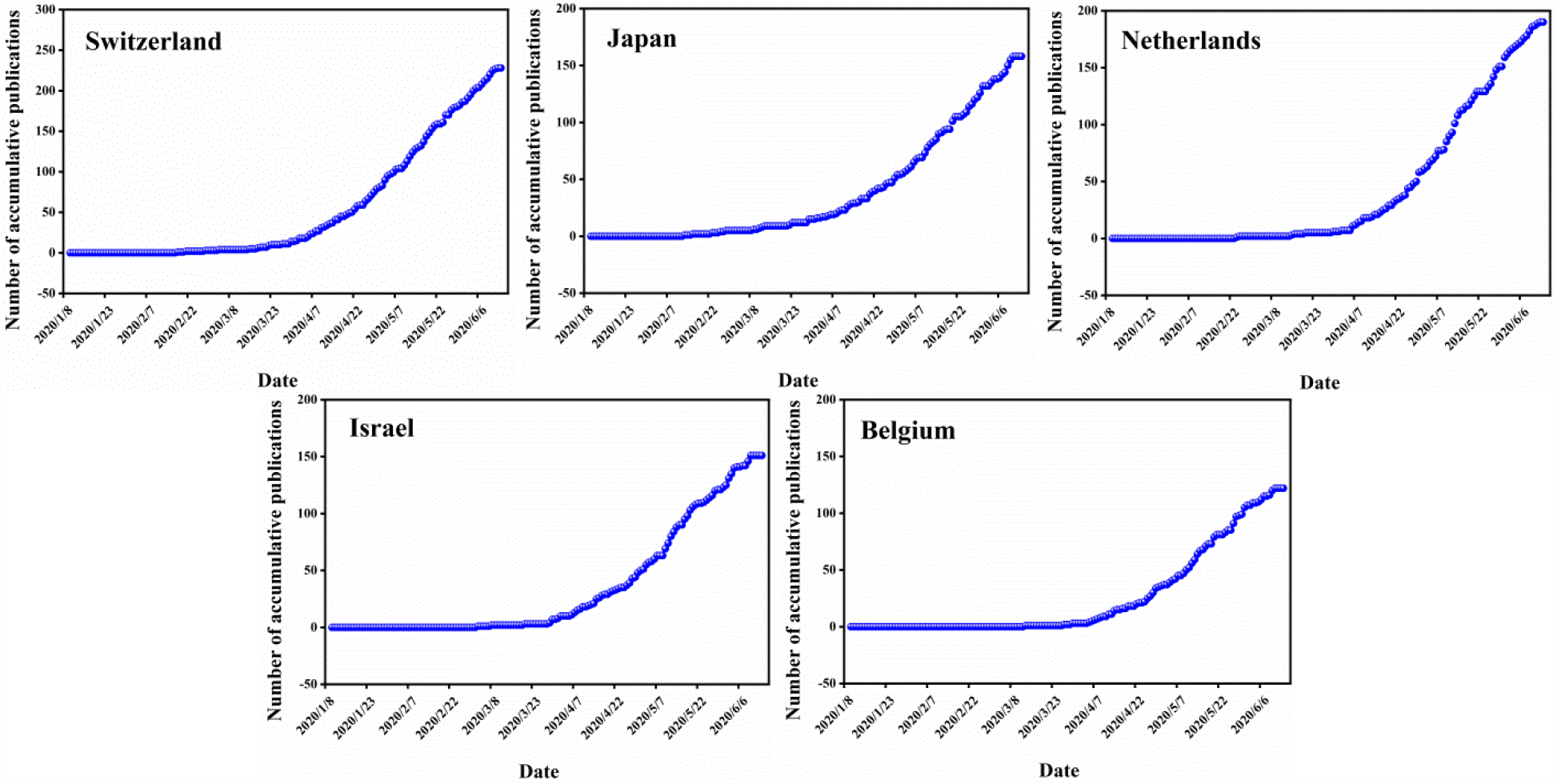
Association between the number of publications and the publication date for each top 20 (i.e., most publications) countries.

**Fig. S2.**
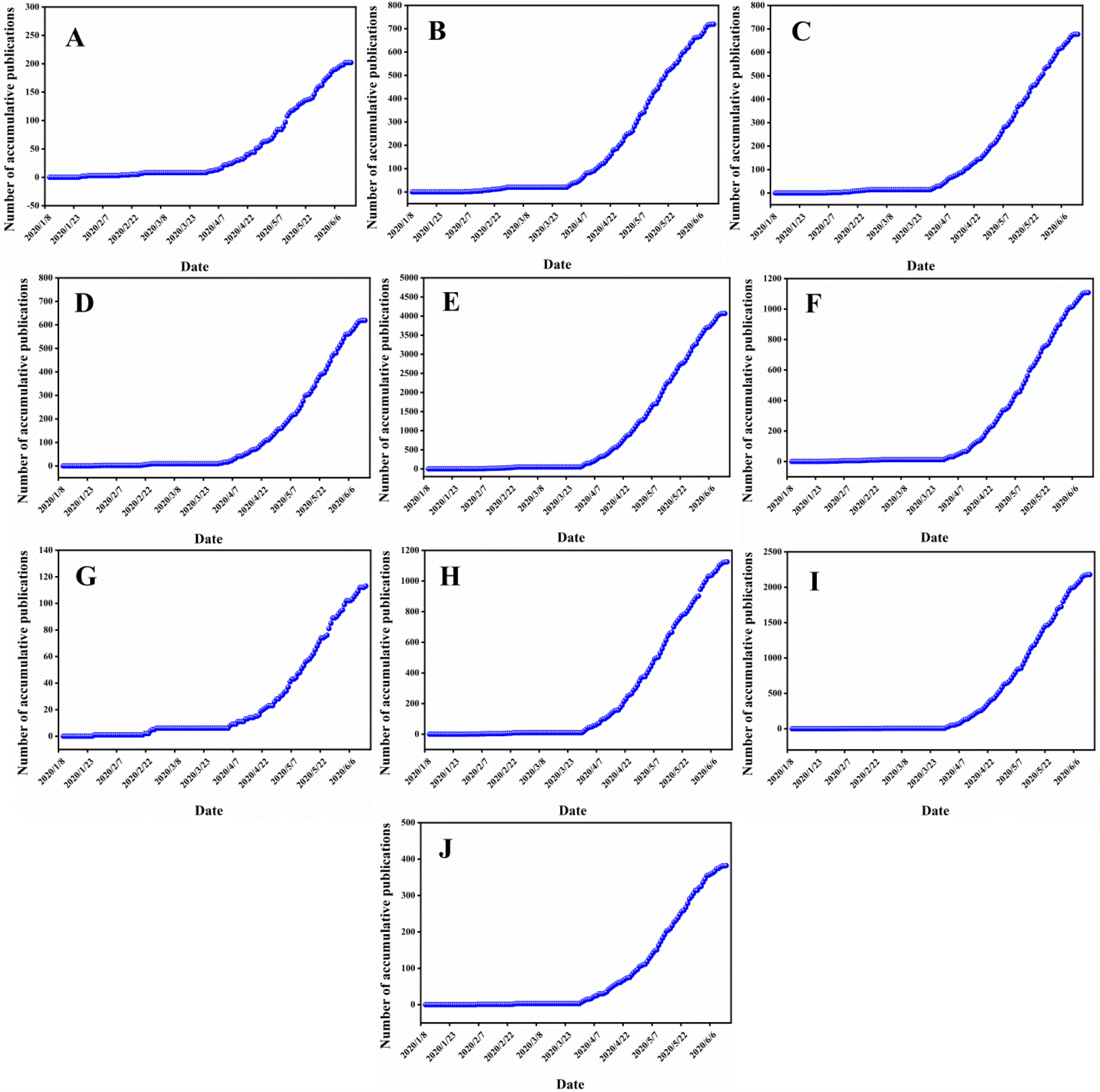
Association between the number of publications and the publication date for each topic, A∼J indicate A. *Origin and gene sequencing*, B. *Epidemiological characteristics*, C. *Clinical characteristics*, D. *Pathology and molecular biology*, E. *Diagnostic methods*, F. *Pharmaceutical treatments*, G. *Vaccine development*, H. *Prevention and control strategies*, I. *Studies targeting a specific population*, and J. *Review*.

**Fig. S3.**
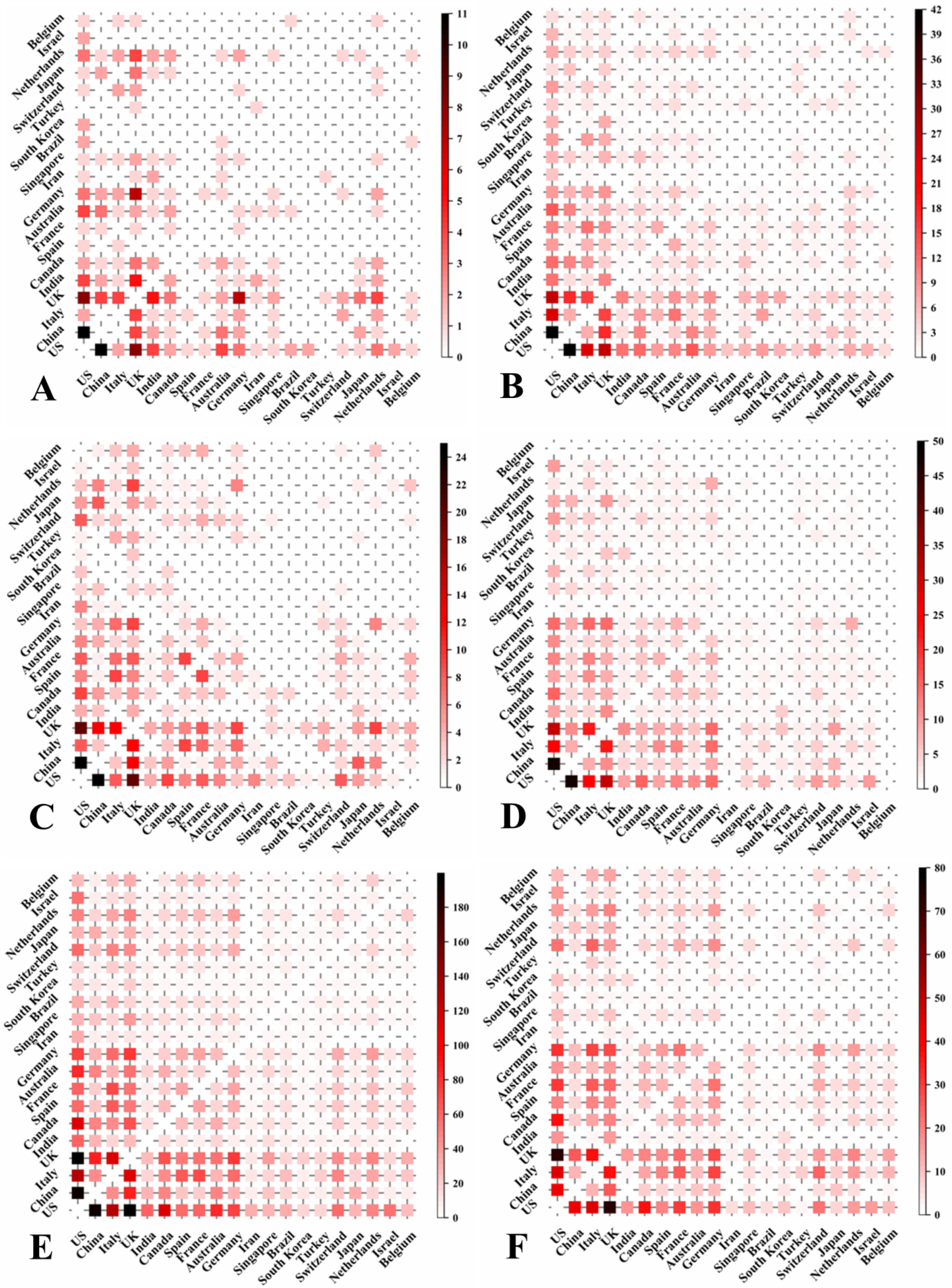
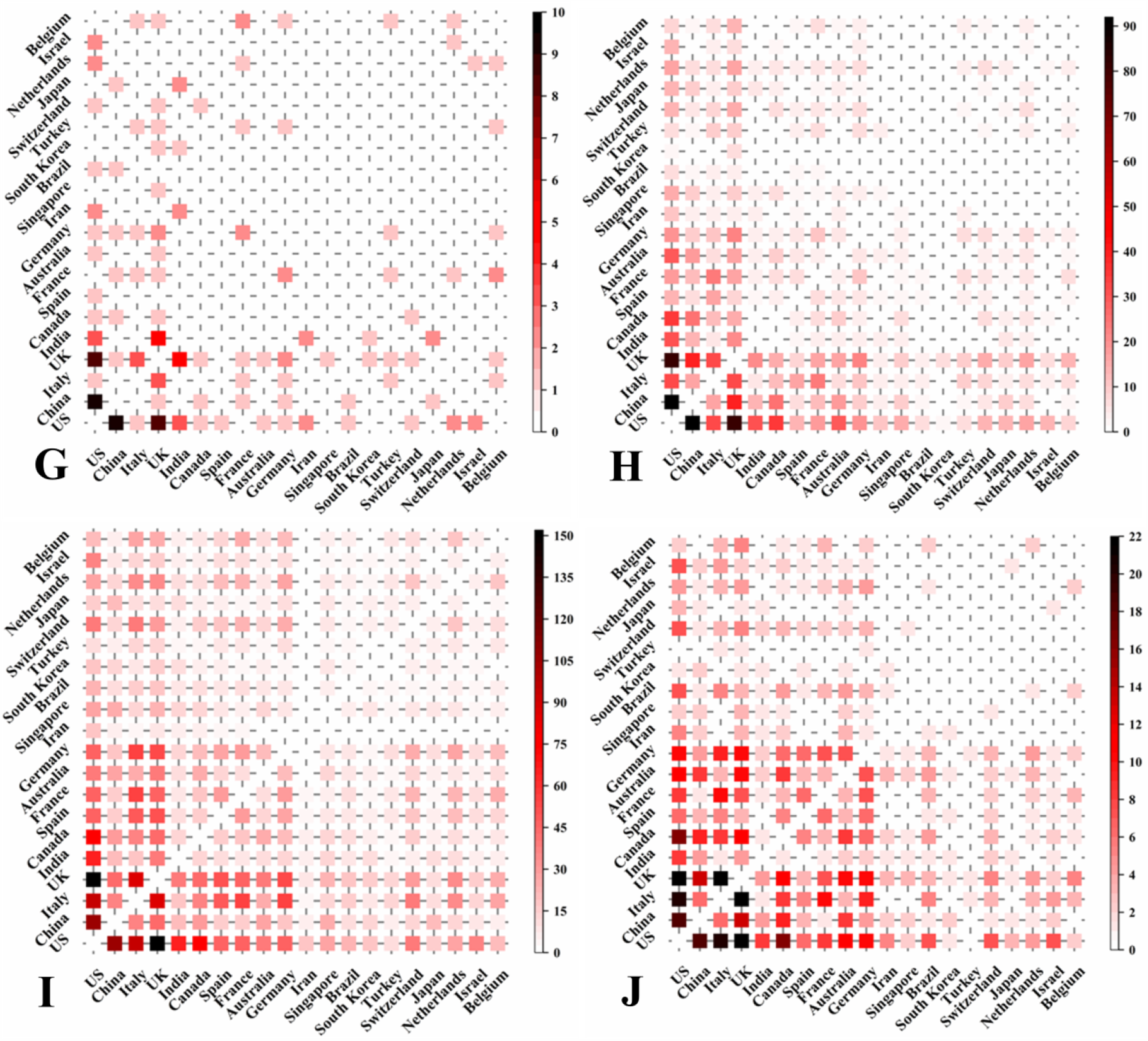
Presentation of cooperation strength between any two top 20 countries (i.e., most publications) on each topic; A∼J indicate A. *Origin and gene sequencing*, B. *Epidemiological characteristics*, C. *Clinical characteristics*, D. *Pathology and molecular biology*, E. *Diagnostic methods*, F. *Pharmaceutical treatments*, G. *Vaccine development*, H. *Prevention and control strategies*, I. *Studies targeting a specific population*, and J. *Review*.

**Table. S1.**
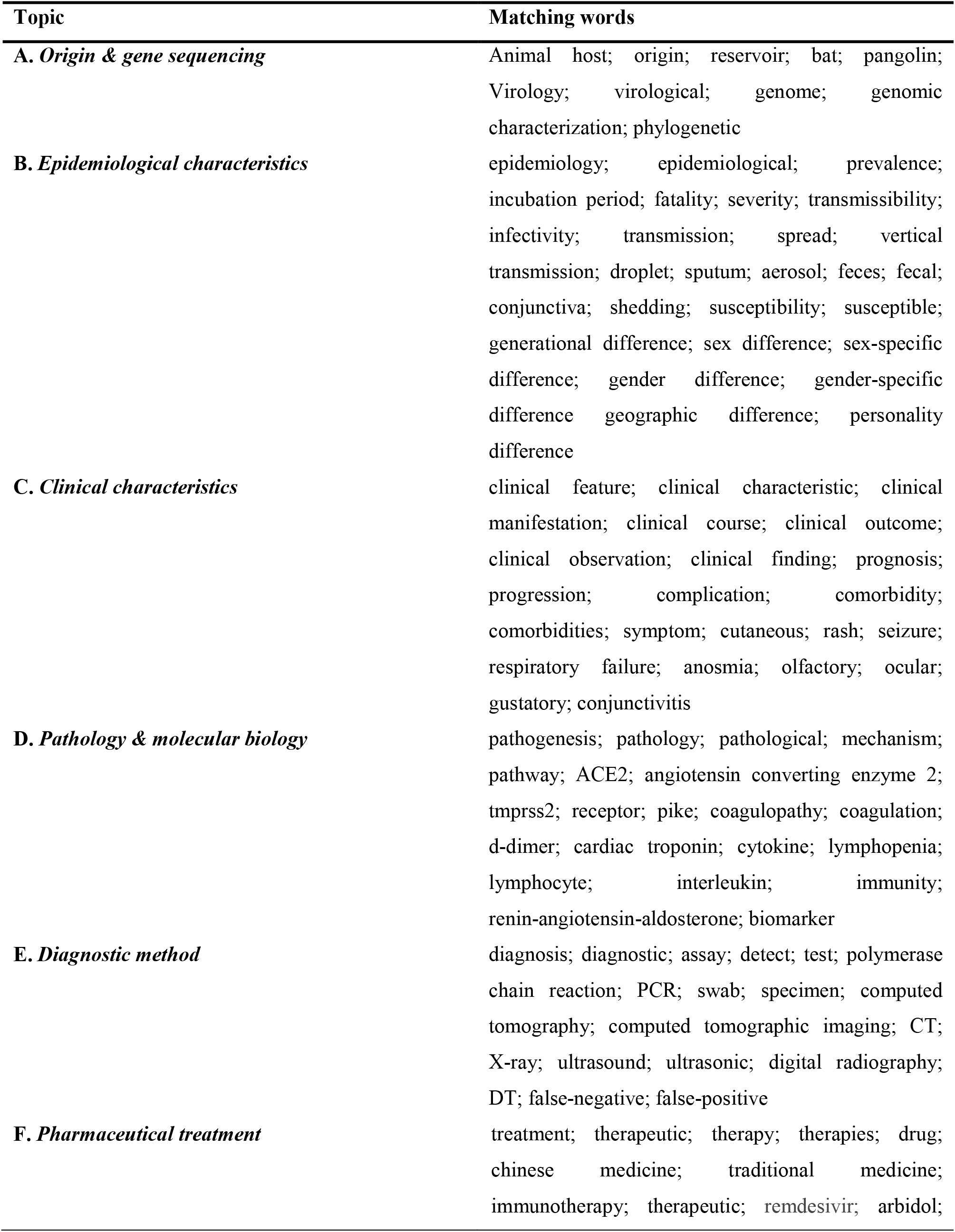
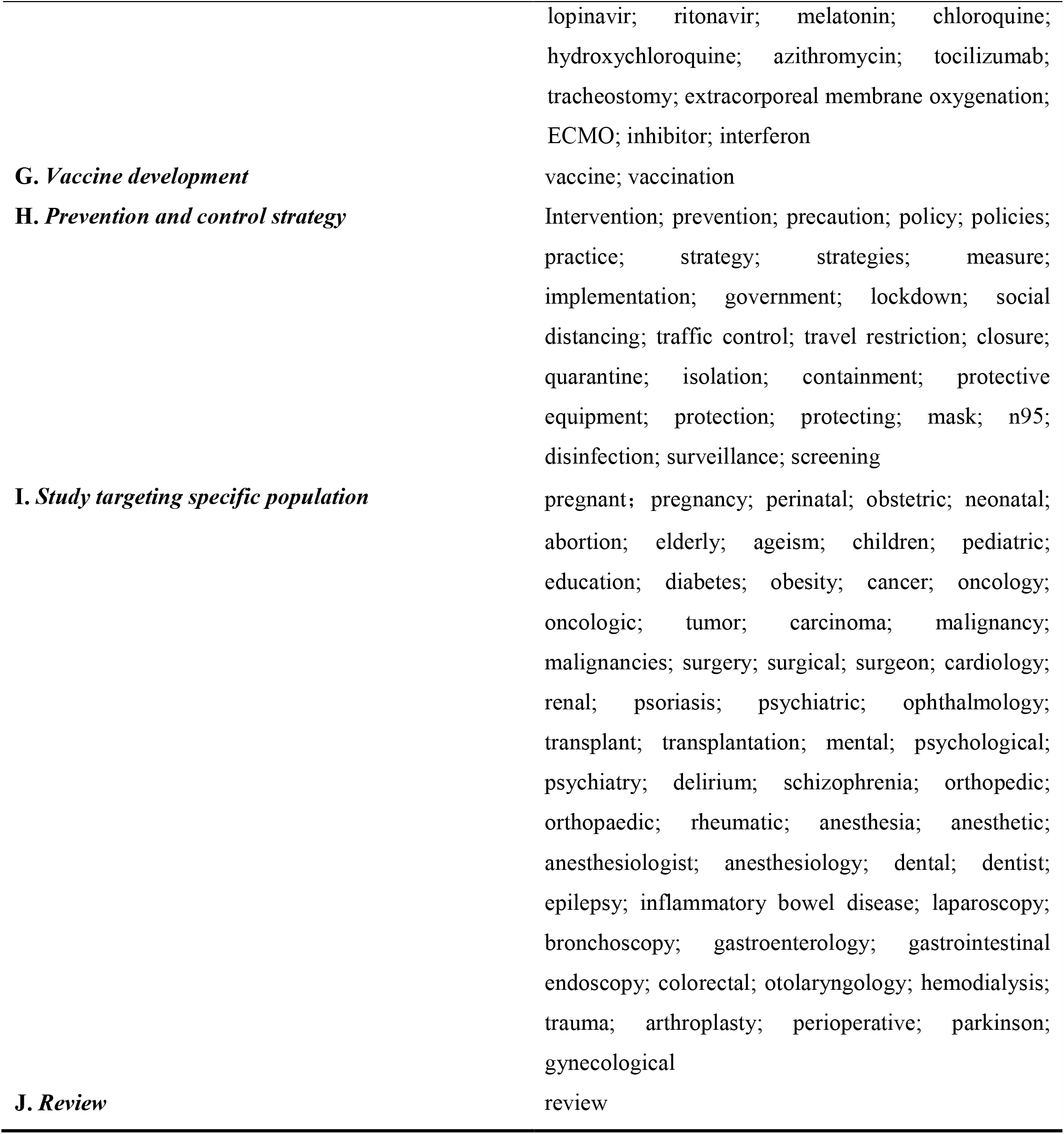
Topic division and the corresponding matching words.

